# Calculation and meaning of “excess mortality”: A comparison of Covid- and pre-Covid mortality data in 31 Eurostat countries from 1965 to 2021

**DOI:** 10.1101/2022.12.22.22283850

**Authors:** Bernhard Gill, Theresa Kehler, Michael Schneider

**Affiliations:** Institute for Sociology, Ludwig-Maximilians-Universitaet Muenchen, Germany

## Abstract

Determining “excess mortality” makes it possible to compare the burden of disasters between countries and over time, and thus also to evaluate the success of mitigation measures. However, the debate on Covid-19 has exposed that calculations of excess mortalities vary considerably depending on the method and its specification. Moreover, it is often unclear what exactly is meant by “excess mortality”. We define excess mortality as the excess over the number of deaths that would have been expected counter-factually, i.e. without the catastrophic event in question. That is, we include all normally occurring flu and heat waves, which are excluded by some authors with the consequence that they almost always record low expected values and correspondingly high excess mortality rates. Based on this definition, we use a very parsimonious calculation method that is easy to understand even for laypersons, namely the linear extrapolation of death figures from previous years to determine the excess mortality during the Covid-19 pandemic. But unlike other literature on this topic, we first evaluated and optimised the specification of our method using a larger historical data set in order to identify and minimise estimation errors and biases. The result shows that the excess mortality rates continuously published by international statistical offices – OECD and Eurostat – are often inflated and would have exhibited considerable excess mortalities in many countries and periods before Covid-19, if this value had already been of public interest at that time. It also reveals that mortality rates already fluctuated strongly in the past and that in a third of the countries studied, individual values from the past exceed the current fluctuations due to the Covid-19 pandemic. Three conclusions can be drawn from this study and its findings: 1) All calculation methods for current figures should first be evaluated against past figures. 2) The definition of excess mortality used should be made explicit. 3) Statistical offices should provide more realistic estimates.

## 1) Introduction

Covid-19 was undoubtedly associated with increased mortality in many countries. But how high was this mortality compared to temporary excess mortality waves in the recent past? “Excess mortality” is a concept that has been known in expert circles since the mid-19th century (Honigsbaum 2020), but Covid-19 has brought it to wider attention even in the general public. It has recently become a means of public alarm, which makes it advisable to take a closer look at the semantic and statistical construction of this term.

Excess mortality is generally defined and determined quite simply: The current mortality is compared with the average mortality figures of the past for the territorial unit and season in question. If the present number of deaths exceeds the average, one speaks of “excess mortality”. This simple demographic method has many advantages: The results are promptly available since there is no need to rely on often uncertain declarations concerning the cause of death. This method has already been used for some time to analyse, for example, the extent of flu epidemics in winter or heat waves in summer without resorting to medical diagnoses (e.g. Canoui-Poitrine et al. 2006; Nunes et al. 2011). Since death itself is a fact that requires little interpretation, excess mortality can also be compared across long periods of time and different cultural groups exclusively on the basis of reasonably reliable demographic statistics.

In the case of Covid-19, tests for detecting an infection were developed very early on. Nevertheless, doubts concerning the daily numbers of Covid-associated deaths persisted: Were sufficient tests available everywhere? How many people died indirectly due to the epidemic, for example due to overcrowded hospitals? To what extent did all those who died with a positive test actually die from the disease or rather because of pre-existing co-morbidities? Since these questions are often difficult to answer, epidemiologists such as Thomas Beaney and colleagues (2020) refer to the excess mortality method as the “gold standard” to assess the extent of the Covid-19 pandemic.

In the context of Covid-19, however, demographic excess mortality was not only treated as a scientific topos in comparatively small specialist circles, but was also received by the general public. Since the onset of the pandemic, national and international statistical agencies such as the OECD and Eurostat have begun publishing excess mortality figures regularly and as quickly as possible. These figures are then frequently disseminated in the mass media. Thus, “excess mortality” has developed from a terminus technicus in limited expert circles to an object of public alarm, which is why increased methodological care is required to avoid misunderstandings. As with other politically relevant thresholds (Schmidt et al. 2008), more thought and discussion is needed to shape the concept of excess mortality.

First of all, it is necessary to clarify what is meant by excess mortality. In the literature, there are two meanings that must be distinguished from each other. In the first meaning, years with particularly low mortality are taken as the starting point for determining excess mortality. For example, the excess mortality of a current influenza wave is then calculated as the difference between the currently increased death figures and the particularly low mortality count in years in which influenza is hardly noticeable (Shkolnikov et al. 2022). The second meaning, and the one most commonly used in the case of Covid-19, was already briefly outlined above: One takes the average mortality of the past (including influenza and all other causes of death) as a basis for estimating an expected value and calculates the excess mortality as a percentage of the difference [1]:

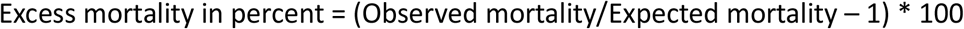

The next question is how to determine the value of the expected mortality. The simplest method has already been indicated above: one takes an average value over a defined number of past years. The OECD’s statistical office, for example, takes the average death counts of the last five years, while Eurostat uses the average of the last four years before Covid-19 as the expected value. However, there is criticism of this method. While averages from the past are suitable for smoothing out random fluctuations between years, they are not able to capture falling or rising trends in mortality figures.

Falling trends may be the result of increasing life expectancy, while rising trends are caused by increasing population numbers, especially in older birth cohorts. In many European countries, we have been observing upward trends in death counts for some time now [2], as is in fact predicted by the theory of the second demographic transition (Lesthaege 2014). If this trend towards rising death rates is not taken into account, excess mortality is regularly overestimated because the average of the past years is lower than the upward trend.

To address this problem, some authors suggest taking into account the varying age cohort composition and their specific mortality rates, thus capturing the trend towards higher mortality rates in ageing populations (e.g. Levitt et al. 2022a; Kowall et al. 2021). Conversely, however, this creates the problem that rising life expectancy is disregarded; the expected values may therefore be too high and excess mortality would be underestimated accordingly (Schöley 2021).

As long as the excess mortality is very significant, as was the case with Covid-19 particularly concerning outbreaks in some local hotspots, a lack of precision in the expected values presents no problem as the estimation error tends to be irrelevant when the number of excess deaths is high. This changes, however, when entire countries are compared on the basis of longer periods – for example, over one or two years – because here the excess mortalities sometimes do not turn out very clearly and even lower than expected mortalities may be recorded (e.g. Sanmarchi et al. 2021). Often, these country comparisons, which are now increasingly emerging in the literature, implicitly or explicitly raise the question: how well did each country cope with the pandemic (Karlinsky/Kobak 2021; Wang et al. 2022; Aburto et al. 2022; Pifarré i Arolas et al. 2022)? This also involves the question of how successful the bundles of measures chosen by the respective governments – restrictive or less restrictive – were (Nepomuceno et al. 2022). Because of the relatively small differences between countries, a higher degree of precision is required here than in the study of individual outbreaks.

It is all the more astonishing that hardly anyone has evaluated their own estimation method so far. The easiest way to do this is to use mortality figures from the past: How well would the mortality figures have been predicted by the method employed for a period before Covid-19, i.e. before the state of emergency? For only when a systematic assessment of the measurement error of the respective method has been carried out on the basis of comparative data can one actually validate the resulting statements on excess mortality during the Covid-19 pandemic. Indeed, several recently published papers show that the figures on excess mortality vary greatly depending on the method and its specification (Schöley 2021, Nepomuceno et al. 2022, Levitt et al. 2022a; Levitt et al. 2022b; Mazzuco 2022; Ferenci 2022). The over- and underestimates are also quite divergent for different countries, so that not only does the overall level rise or fall, but also the ranking of countries changes with the method – i.e. individual countries may do “well” with one method and “poorly” with another.

Levitt and colleagues (2022a) compare four prominent methods as well as their own age-stratified model with the finding that in the 33 countries studied that have relatively reliable population statistics, the excess mortality of all these countries together is between 1.6 and 2.8 million deaths, in the highest case almost twice the lowest estimate. For individual countries like Germany, the deviation is even more pronounced – the lowest estimate here results in 55,000, the highest in 203,000 additional deaths [3]. In general, the relevant studies also emphasise the enormous sensitivity of most methods to the time frame chosen to calculate the expected values (Levitt et al. 2022b, Nepomuceno et al. 2022, Mazzuco 2022). In this context, Ferenci (2022) refers to the significance of the strong influenza wave in 2015. Since most studies for determining the expected value for the Covid 19 pandemic refer to the five preceding years, i.e. 2015 to 2019, the high number of deaths in 2015 and the low number of deaths in 2019 result in a trend that is relatively flat or – in the original and later corrected spline model of the WHO for Germany – even declining due to these coincidences. In consequence, the expected value for 2020 and 2021 is estimated significantly too low (Ferenci 2022, Figure 1).

In contrast to other alarm values (Schmidt et al. 2008), it is also noticeable that threshold values are rarely given. These could serve to distinguish normal excess mortalities from extraordinary and therefore politically really noteworthy excess mortalities. First of all, it should be noted that the term “excess mortality” in everyday language automatically implies the idea of an exceptional event, although according to the definition used by experts a certain excess mortality is completely normal, because the fluctuations of excess mortality and lower than expected mortality would balance out to zero over the years, provided that the expected value was estimated correctly. So, in order to determine whether excess mortality is exceptional for a particular country in a particular year, one has to study the figures from the past: For example, is an excess mortality of 2.5 percent in Germany in the Covid year 2020 exceptional, i.e. greater than most excess mortality rates in the last 10 or 50 years?

In conclusion, methods for determining excess mortality have rarely been evaluated and compared on the basis of actual and longer-term mortality figures. The present article attempts to fill this gap. A computation method for the expected value is proposed that takes into account both fluctuations and trends while being very simple to calculate, correspondingly easy to understand and parsimonious in its data requirements. The method described is then tested for accuracy using mortality data from 31 Eurostat countries for the period from 1965 to 2019. It is shown that it can compete in accuracy with very complex methods and performs significantly better than the calculation methods used by the OECD and Eurostat. Figures for excess mortality for 2020 and 2021 are then derived and discussed with respect to historic excess mortality between 1965 and 2019. This reveals that the excess mortality of the Covid-19 years is within normal fluctuations in 10 countries while it exceeds extreme values of the examined time frame since 1965 in 21 countries. The discussion concludes with two policy recommendations: First, statistical offices could use better estimation methods without calculations becoming too complex and thus suspected of being non-transparent. Second, as with other warnings, appropriate thresholds should be set. According to our findings, alert for excess mortality in the past would have been quite normal with the calculation methods currently used by statistical offices – that is, if excess mortality had already been reported in the period before Covid-19.

## 2) Method development

As the central basis of our calculations, we use the annual death figures of all 31 countries in Eurostat which provide complete data for the period from 1960 to 2020 (code: demo_magec) [4]. The values for 2021 were calculated from the more recent (and therefore provisional) data of weekly death counts in Eurostat (code: demo_r_mwk_ts). To compensate for distortions due to leap years, the death figures in these years were multiplied by a correction factor (1/366*365) and thus all years were normalised to 365 days.

The method for estimating the expected values is based on the considerations that there are annual fluctuations and longer-term upward and downward movements, as already indicated above. Figure 1a shows the situation before the pandemic in the years 1960 to 2019, using Germany as an example. It can be seen that the number of deaths tended to rise until the mid-1970s, then fell until the mid-2000s and has since been rising again.

**Figure 1a:**
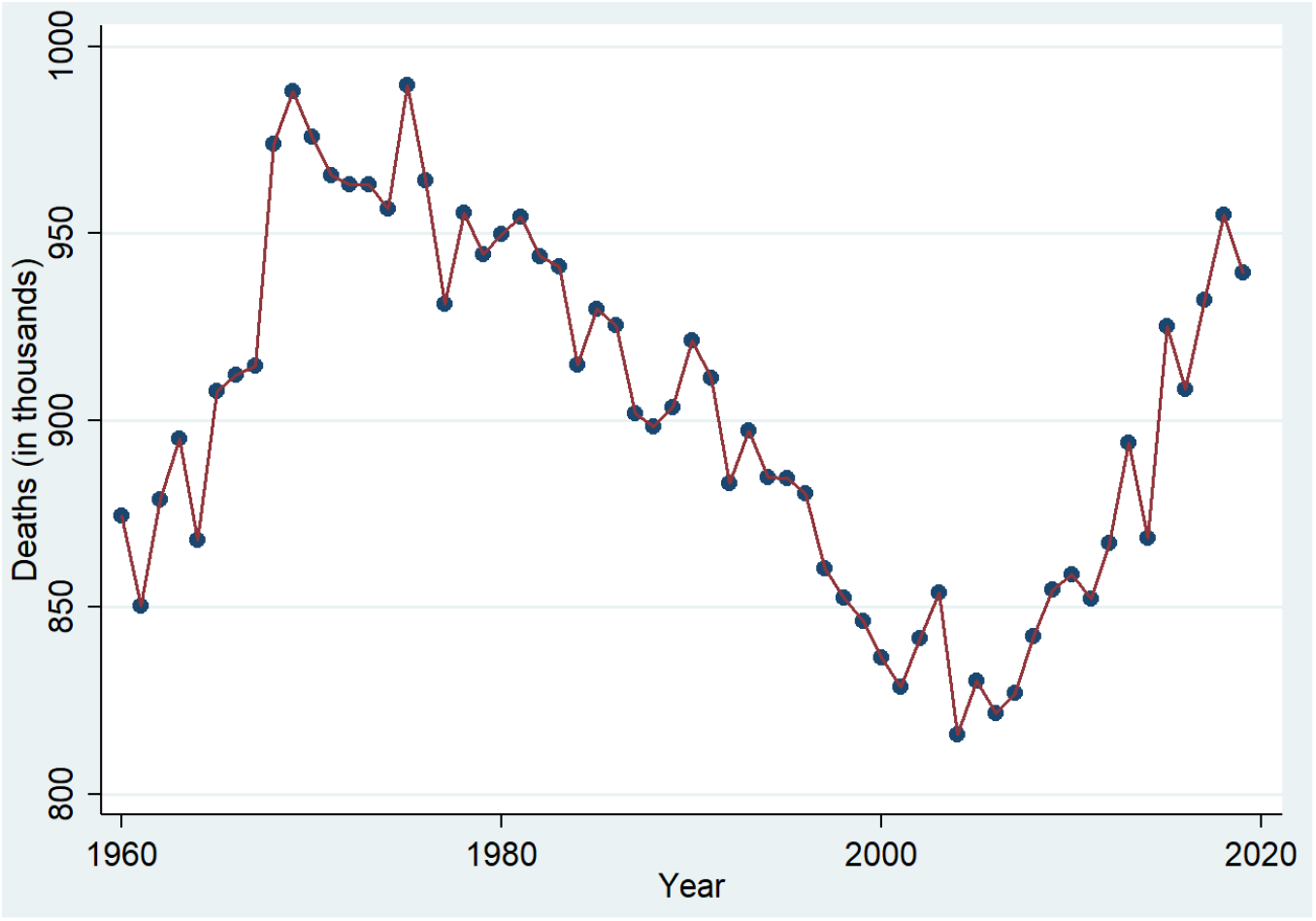
Annual number of registered deaths in Germany before Covid-19.

To capture longer-term tendencies and smooth out year-to-year fluctuations, a linear trend is estimated using the death count in a specified number (n) of previous years:

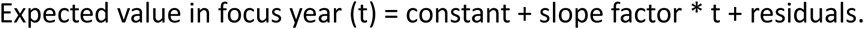

The constant and slope are determined by means of the most common method, Ordinary Least Squares (OLS) regression, which is also implemented in simple spreadsheet programs, i.e. in widely available standard software. How many previous years are included in the estimation is theoretically a question of balance: the more years are included, the better random fluctuations between the years are evened out, but the slower the expected value reacts to changes in the trend. Hence, in the following it is empirically determined which number (n) of previous years provides the most accurate estimate and is therefore recommended.

But how is the quality of the expected value to be assessed[5]? First, the **M**ean **A**bsolute **P**ercentage **E**rror (MAPE) of the estimation procedure must be determined. To do this, the absolute value of the respective deviations of the observed value from the expected value is added up and divided by the number of corresponding years (1965 to 2019 = 55 years):

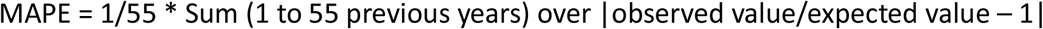

This first parameter should be relatively low; but it will be a few percentage points above zero, since the annual fluctuations resulting mainly from the occurrence or absence of more severe influenza waves in winter and heat waves in summer cannot be predicted. Even if an “ideal value” can hardly be obtained here, the relative performance of an estimation method compared to another estimation method can nevertheless be easily identified on the basis of the average level of the estimation error (MAPE): The lower the estimation error, the better the method.

Secondly, the predominant direction of the estimation error, its bias, must be determined. The calculation is similar; but instead of the absolute value, the actual values are summed in the conventional way so that positive and negative deviations balance each other out. However, only the last five years before the focussed event are considered (for Covid-19: 2015 to 2019), because this shorter period is more relevant for the present than the longer period, in which the bias levels out if the trend is reversed:

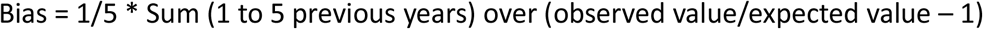

Ideally, this second parameter should be as close to zero as possible. A positive bias means that the expected value tends to underestimate the observed value and consequently, on average, shows inflated excess mortality rates. The reverse is true for a negative bias; in this case, excess mortality is underestimated.

Figure 1b compares two methods for forming the expected value based on the death figures in Germany. It can be seen that the five-year mean, as used by the OECD, always lags behind the actual development and estimates the expected values too low when the numbers are rising and, conversely, too high when the mortality is declining. In contrast, the 5-year trend is much closer to the actual development. This is also reflected in the two performance indicators. MAPE (the absolute estimation error) is 1.78 percent on average for the 5-year trend, but 2.29 percent for the 5-year mean; the bias is 0.03 percent and 0.27 percent respectively. The differences become even clearer if either ascending or descending phases are considered. For example, taking only the years 2010 to 2019, MAPE is 1.96 percent for the trend method and 3.34 percent for the mean method; the bias is -0.03 percent for the former and 3.34 percent for the latter. Thus, in phases with a constantly increasing or decreasing tendency, the bias in the OECD mean method is very pronounced.

**Figure 1b:**
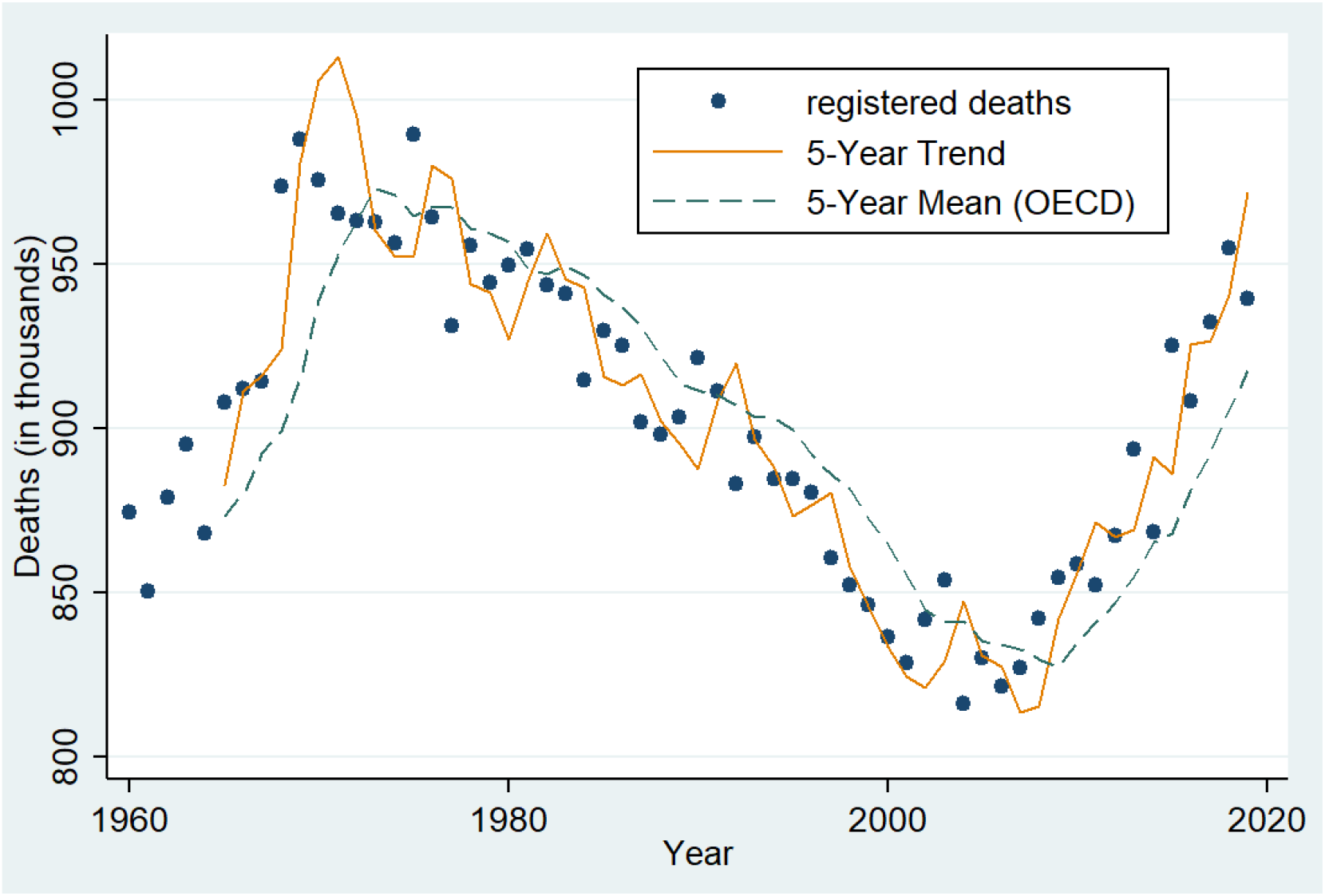
Annual number of deaths in Germany from 1960 to 2019 with two different expected value forecasts.

## 3) Results

First, we tried to find out which trend has the most favourable performance indicators. Table 1 shows that, calculated over all countries and years (1960 to 2019), the estimation error (MAPE) is lowest for the 7-year trend. In contrast, the 3-year trend has the lowest bias. In this respect, the 5-year trend represents a good compromise between the two performance indicators – accordingly, our further calculations are based on this choice. If we compare the 5-year trend with the standard method of statistical offices, the 5-year or 4-year mean, we see a particularly marked difference in terms of bias; this is 9 times higher for the 5-year mean, i.e. the OECD method, and 7 times higher for the 4-year mean, the Eurostat method, than for our 5-year trend. The difference in the estimation errors (MAPE) is not as drastic and amounts to 22 and 11 percent.

**Table 1:** Comparison of estimation errors (MAPE) and bias of different methods for 31 nations. The most favourable values are highlighted in bold. The 5-year trend thus represents the best compromise (marked yellow).

| Method | MAPE (1960 to 2019 mean) | Bias (2015 to 2019 mean) |
| --- | --- | --- |
| 2Y-Trend | 4,54 % | 0,23 % |
| 3Y-Trend | 3,33 % | **0,20 %** |
| 4Y-Trend | 2,90 % | 0,29 % |
| 5Y-Trend | **2,77 %** | **0,32 %** |
| 6Y-Trend | 2,71 % | 0,56 % |
| 7Y-Trend | **2,69 %** | 0,78 % |
| 8Y-Trend | 2,70 % | 1,01 % |
| 9Y-Trend | 2,72 % | 1,24 % |
| 4Y-Mean | 3,07 % | 2,43 % |
| 5Y-Mean | 3,38 % | 2,86 % |

In the following, we examine how the use of the 5-year trend, i.e. the method proposed here, affects the estimate of excess mortality in the Covid 19 period, i.e. for the years 2020 and 2021 together. For simplicity, only the OECD method (the 5-year mean) is now used for comparison, as the Eurostat method (the 4-year mean) would yield very similar results. For the second pandemic year (2021), we have used the trend or mean from 2015 to 2019 to estimate the expected value so that the calculations are not biased by the increased number of deaths in the first pandemic year (2020). Figure 2 shows the average annual excess mortality over the two-year period 2020 to 2021, ordered from left to right using the excess mortality values by country derived from the 5-year trend. The differences between the two methods are very pronounced for some countries. They result from the fact that the mean method, as already mentioned, does not take into account underlying gradients: If the death figures show an increasing tendency, as is the case in Northern and Western Europe, the mean method overestimates excess mortality; if they show a decreasing tendency, as is partly the case in Eastern Europe, this leads to an underestimation (cf. also the changing gradients in Figure 1 above) [6].

**Figure 2:**
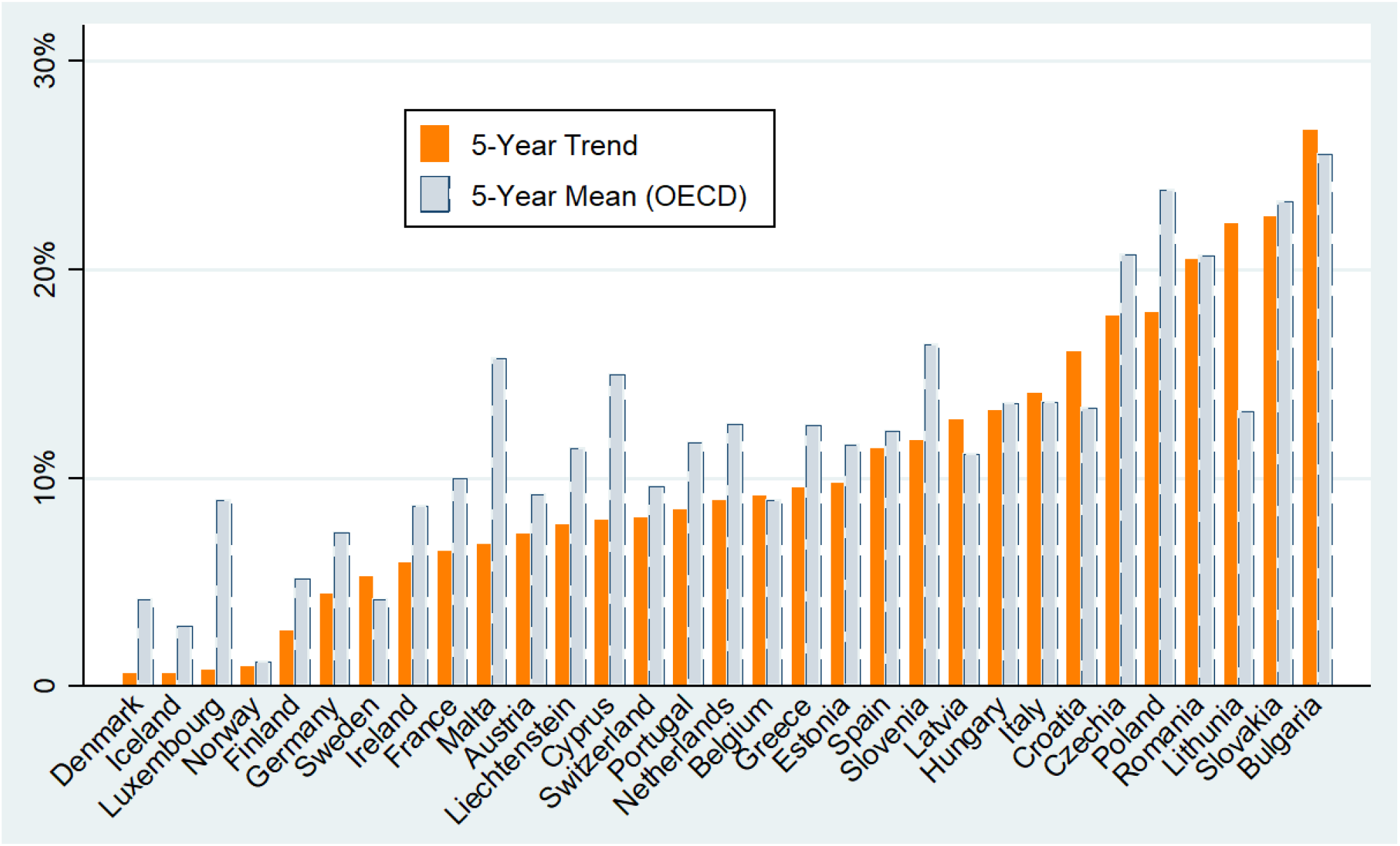
Average annual excess mortality in the two-year period 2020 to 2021, i.e. during the Covid-19 pandemic. We compare our proposed method (5-year trend) with the estimates of the OECD statistical office (5-year mean).

It can also be seen in Figure 2 that excess mortality during the pandemic in Europe shows a high variance – ranging from 0.8% in Denmark to 27% in Bulgaria. These are the currently much-noticed differences between countries, which are often spontaneously attributed to the measures against the pandemic and their more or less strict compliance. In Germany for example, the rather mild excess mortality is usually assigned to the heavy restrictions during the pandemic while Sweden was criticised for its liberal approach. But in hindsight, we see Germany and Sweden more or less on par, in the 6^th^ and 7^th^ position. In general, it becomes obvious that the lowest excess mortality is predominantly found in Northern Europe, followed by Western Europe, Southern Europe and Eastern Europe, thus clearly following a negative socio-economic gradient. The correlation coefficient between excess mortality and wealth is indeed quite strong (in the negative direction) and highly significant [7].

However, we do not know how high the variations in excess mortality were in the past and how much they differed between the countries. In other words: Before we try to interpret death tolls and containment measures during the pandemic, we should analyse the normal mortality fluctuations from pre-pandemic years. Figure 3 attempts to answer this question by comparing the average annual excess mortality during the Covid-19 pandemic with the excess and the lower than expected mortality from 1965 to 2019. Here it becomes visible that the mortality figures in countries with smaller populations – due to the law of small numbers, i.e. for statistical reasons – fluctuate strongly anyways. This applies in particular to Liechtenstein (38,000 inhabitants), Iceland (335,000), Malta (430,000) and Luxembourg (583,000). However, even in countries with a comparatively large population, such as Germany, France, Italy and Spain, high fluctuations of plus/minus five percent are relatively normal. In this respect, it can be stated that in 10 out of 31 countries the average annual excess mortalities in the period from 2020 to 2021 do not exceed the range of excess mortalities in the period from 1965 to 2019. Furthermore, it is noteworthy that the highest excess mortality during this period does not occur in or around any particular year – i.e. it cannot be explained by other pandemics, such as the Hong Kong flu, which spread worldwide from 1968 to 1970 (Viboud et al. 2005).

**Figure 3:**
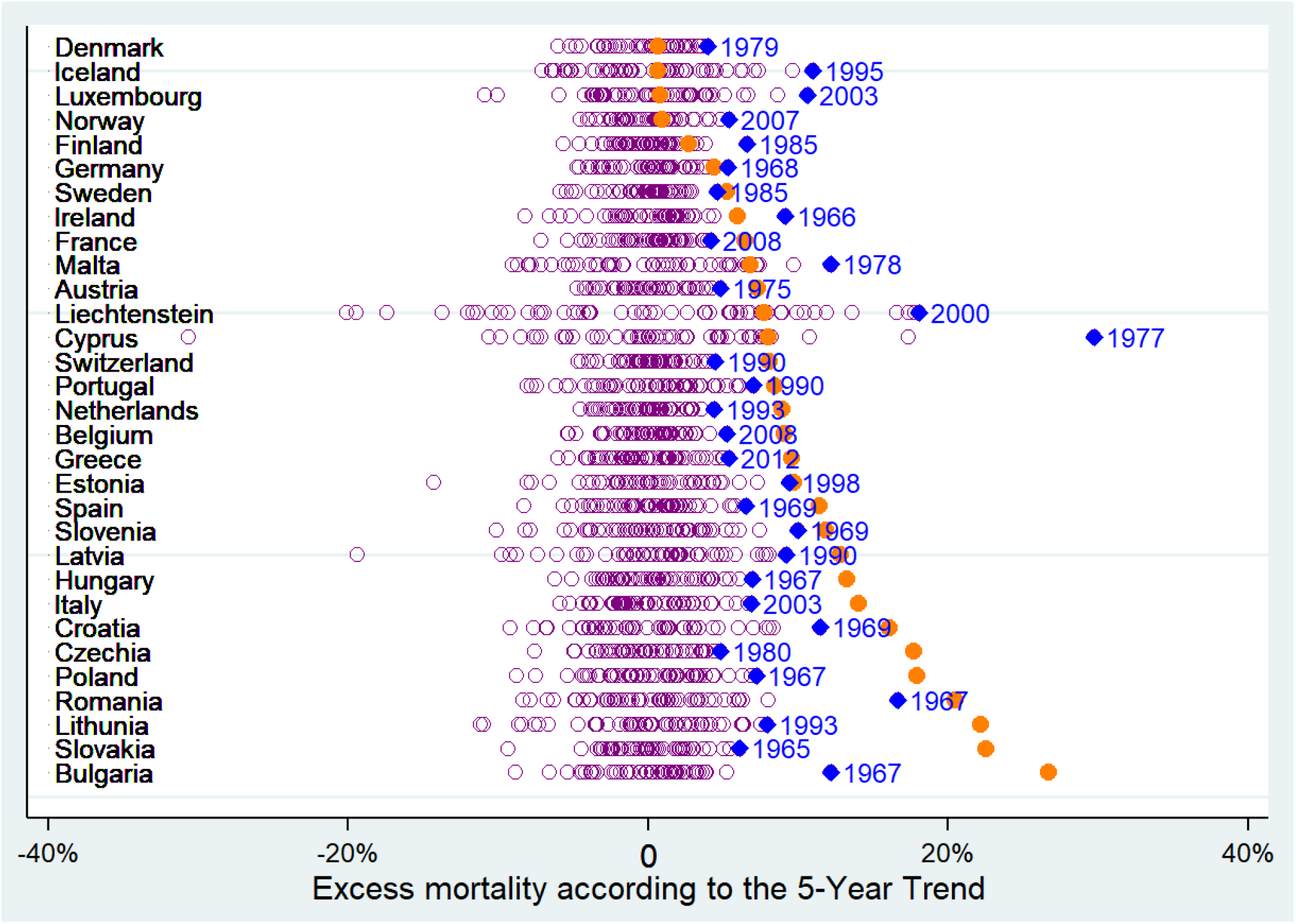
Average annual excess mortality during the Covid-19 pandemic (orange dots) compared with excess mortality values from 1965 to 2019 (purple rings). Blue diamonds with year labels indicate the respective year with the highest excess mortality in the pre-Covid-19 period.

## 4) Discussion and conclusions

Three main results should be noted and discussed: First, the importance of the term “excess mortality”; second, the need for an evaluation method; third, the accuracy of our calculation method compared to other methods commonly used in the literature.

Firstly, it has been shown that relatively high variations in death figures, and thus in excess mortality, already occurred in the past. Our estimation procedure also reveals years with remarkable lower than expected mortality. In the light of these fluctuations, the question arises as to which horizons of meaning should be invoked with the term “excess mortality”. Three different notions are conceivable and should not be confused with each other: a) The one established by William Farr (Honigsbaum 2020), used in this text and still most common among statistical offices today, is that of a deviation from a longer-term average or trend, which is understood as “demographic normality”. A threshold value is not envisaged here. b) In contrast, the epidemiological and demographic literature often takes the view that all “preventable” deaths should be understood as excess mortality, i.e. also “normal” flu deaths, heat deaths, traffic victims, etc. (e.g. Nielsen et al. 2019; Shkolnikov et al. 2022). As a result, the baseline from which excess mortality is calculated is generally drawn lower and at the same time the normative reference point is shifted. The benchmark is no longer the socially accepted normality before the exceptional event under consideration, but the author’s ideas about what he or she considers avoidable and what not. While these ideas may be plausible, they are moving targets in scientific debates and not fixed points of reference across time, space, and culture, that could be used to determine, for example, the magnitude of a pandemic on a historical scale (Gill et al 2022). c) Finally, there is the possible and, in the public mind, perhaps also (mis-)understood meaning of an alert threshold, which is supposed to indicate an extraordinary event. Here, too, the point of reference is demographic normality, except that it is not the zero point of normal fluctuations between excess mortality and lower than expected mortality that would be taken as the baseline, but an upper threshold value – for example, a threshold value of five percent for the annual values of more populous countries [9]. Assuming this threshold, for Covid-19 no alarm would be triggered in 7 out of the 31 countries considered (for excess mortality in the two-year period).

Secondly, it has been demonstrated that different calculation methods lead to quite different results. It is therefore necessary to calibrate the calculation method used against a past that is considered normal before applying it to an exceptional event. The prevailing neglect of this necessity is all the more astonishing as the methodological reviews cited above show considerable differences in results due to the choice of method. However, considering that excess mortality rates are relevant to public controversies and thus politically sensitive, one should a priori choose a transparent and easily comprehensible method in order to avoid the suspicion of “cherry-picking” – and corresponding alarmist or anti-alarmist motives. Most of the recently published reviews that refer to the problem of method selection remain rather vague in their recommendations for remedial action. Nepomuceno and colleagues (2022), for example, continue to leave the decision on method to the discretion of the researcher: “Moreover, the method used and the reference period (which may or may not include leap week years) should both be chosen carefully.” (p. 21) Only Jonas Schöley (2021) uses past data – similar to ours – to assess the accuracy of different methods, but unfortunately in a way that the results are not directly comparable to ours [9].

Thirdly, we have proposed and tested a calculation method with comparatively good results. We were able to demonstrate that it has a significantly lower bias and higher accuracy compared to the methods used by Eurostat and the OECD. Importantly, this is achieved without higher demands on data availability and mathematical understanding, unlike other methods described in the literature (e.g. WHO 2022). We therefore believe that it represents a good compromise between ease of verification and statistical accuracy. In particular, the bias is very low and seems to be competitive with computationally much more sophisticated methods (the data in Schöley 2021, Figure 4, seem to indicate this, even if they are not directly comparable).

Concerning the deviation of expected from observed values, the accuracy can perhaps still be improved; the estimation error (MAPE) could possibly be diminished: Here it would be particularly interesting to find out whether there is a certain systematics in the fluctuations over the years, as could be revealed, for example, with an ARIMA-procedure commonly used in econometrics for overlapping waves and trends (Shumway/Stoffer 2017). In terms of causality, it would also be worth considering how the remarkably lower than expected mortalities shown in Figures 1 and 3 come about [10]. In the epidemiological literature, mortality displacement effects are occasionally reported, i.e. that in the case of heat or flu waves, severely pre-weakened persons die and therefore lower than expected mortalities can occur afterwards (Armstrong et al. 2017). To the best of our knowledge, these effects have never been studied in larger demographic contexts over longer periods of time.

However, all methods for assessing excess mortality should be evaluated with regard to their estimation error and bias using standardised procedures, in order to be able to exploit their advantages over other measurement methods (such as direct disease diagnosis). This is particularly important since pandemics – throughout history (Cohn 2012) – have generated considerable controversy and therefore any appearance of non-transparency and political bias on the part of science should be avoided as far as possible. Particularly the methods of the statistical offices – as OECD and Eurostat – are in urgent need of improvement due to their high bias and broad impact. Excess mortality as a summary indicator of a pandemic crisis is too relevant for health policies and public health interventions to risk potentially misleading calculations and interpretations.

## Data Availability

The spreadsheet file with the respective data and computations is made public on OSF:
https://osf.io/5y4sd/files/osfstorage/63a47dc17b5c800255227a92

## 5) Funding

The study was partly supported by the Volkswagen Foundation, grant Az. 99 026.

## 6) Competing interest

None of the authors is aware of any competing interest.

## 7) Data availability

The spreadsheet file with the respective data and computations is made public on https://osf.io/5y4sd/files/osfstorage/63a47dc17b5c800255227a92

## Notes

### Competing Interest Statement

The authors have declared no competing interest.

